## Supplementary Appendix for "Characteristics of individuals who received a complete, 2-dose mpox vaccine regimen as part of the public health response to the mpox epidemic in Ontario, Canada: A CIRN study"

### **Supplementary Methods**

#### **Linked data repositories**

Ontario has a single-payer health system that provides universal access to physician and hospital services and laboratory testing for all eligible Ontario residents. For this study, we used population-based databases that capture relevant information within this single-payer system.

MVA-BN was administered as pre-exposure prophylaxis to eligible high-risk groups by local public health units at mass immunization clinics in coordination with community-based organizations, with a smaller number administered via physician clinics. The mass immunization clinics were implemented in drop-in settings, including community-based organizations, with a low-barrier approach to accelerate uptake and maximize access. The low-barrier approach meant that confirmation of eligibility criteria was not required, and any individual who came to the mpox-specific mass immunization clinic could receive the vaccine.

The provincial Digital Health Immunization Repository (DHIR) contains individual-level vaccination data (e.g., product, dose, site, date) entered at the time of administration. Data on post-exposure prophylaxis doses administered by primary care providers were submitted to local public health units for entry into DHIR retrospectively.

All mpox specimens collected by health care providers were submitted to Public Health Ontario laboratories for testing. Laboratory results and demographic and clinical data from the test requisition were obtained from LabWare, Public Health Ontario’s laboratory information system.

Data on syphilis testing were obtained from LabWare and the Ontario Laboratories Information System (OLIS), a provincial repository of laboratory testing results from hospital laboratories, community-based commercial laboratories, and public health laboratories. Data on prior diagnoses of bacterial sexually transmitted infections (gonorrhea, chlamydia, and syphilis) were obtained from the integrated Public Health Information System (iPHIS), an information system for the reporting and surveillance of Diseases of Public Health Significance.

History of HIV diagnosis was ascertained from the ICES-derived HIV Cohort, which uses a validated algorithm based on physician office visits with HIV as the diagnostic code to identify individuals living with HIV.^1^ Additionally, HIV pre-exposure prophylaxis prescription data were obtained from the Ontario Drug Benefit (ODB) database, which contains information on all publicly-funded prescriptions for: adults aged ≥65 years; recipients of professional home services and social assistance; children and young adults aged ≤24 years and not covered by a private insurance plan; and recipients of the Trillium Drug program, which helps Ontarians who have high prescription drug costs relative to household income.^2,3^

Information on COVID-19 vaccination was obtained from Ontario’s centralized province-wide COVID-19 vaccine registry, COVaxON. Data on influenza vaccines and other publicly funded vaccines received in physician offices were extracted from the Ontario Health Insurance Plan (OHIP) physician billing claims database, while information on influenza vaccines received in pharmacies was extracted from the ODB database. Data on number of physician office visits and whether individuals had a primary care physician were also obtained from the OHIP physician billing claims database. Data on immunocompromised status was obtained from CCI procedure codes and OHIP feecodes, ODB to identify immunosuppressive medications, the OCR for cancer diagnoses, and Discharge Abstract Database (DAD), Same Day Surgery (SDS), OHIP, and National Ambulatory Care Reporting System (NACRS) for other healthcare encounters that suggested individuals had disorders of the immune system.

We obtained age, sex, postal code, and neighborhood-level income and visible minority quintile from the Ontario Registered Persons Database (RPDB), a population registry with demographic information for all Ontarians with provincial health insurance. We determined the geographic region (public health unit, which we then collapsed into a smaller number of regions) using the postal code and Statistics Canada Postal Code Conversion File plus (version 7B). We determined immigration status using the Immigration, Refugees and Citizenship Canada (IRCC) Database, which includes administrative information related to temporary and permanent residents in Canada.

De-identified data extracted from repositories held at Public Health Ontario (Panorama, Labware, iPHIS) were securely shared with ICES, a not-for-profit research institution, and linked to health administrative databases held at ICES using unique encoded identifiers. Linked data were analyzed at ICES.

### **Table S1.** Definitions of variables used.

| **Variable** | **Definition** |
| --- | --- |
| Laboratory-confirmed mpox infection (used for exclusion criteria) | First positive or intermediate test result on real time polymerase-chain-reaction (PCR) testing for orthopox/mpox on samples collected from any body site. |
| Reason for immunization | Immunization provided for pre-exposure prophylaxis or post-exposure prophylaxis. |
| Number of days between 1^st^ and 2^nd^ dose of MVA-BN | Days between a patient’s first MVA-BN dose and second MVA-BN dose. |
| First dose before or on/after September 30, 2023 | Indicator of whether a patient had their first MVA-BN dose prior to September 30, 2023 or on/after September 30, 2023. |
| Age-group | Age at dose 1 was determined from the Registered Persons Database, and categorized into the following groups: 0-17 years, 18-24 years, 25-29 years, 30-39 years, 40-49 years, 50-59 years, and ≥60 years. |
| Geographic region (public health unit region) | Ascertained from Public Health Unit (PHU) information using postal code of residence as recorded in the Registered Persons Database and Statistics Canada Postal Code Conversion File Plus (version 7B). Regions were defined as follows:  Toronto: PHU 95 (City of Toronto Health Unit)  Peel/York/Durham/Halton: PHU 53 (Peel Regional Health Unit), 70 (York Regional Health Unit), 30 (Durham Regional Health Unit), 36 (Halton Regional Health Unit)  Hamilton/Niagara/London/Windsor: 37 (City of Hamilton Health Unit), 46 (Niagara Regional Area Health Unit), 44 (Middlesex-London Health Unit), 68 (Windsor-Essex County Health Unit)  Ottawa: PHU 51 (City of Ottawa Health Unit)  Rest of Ontario: PHU 35 (Haliburton, Kawartha, Pine Ridge District Health Unit), 55 (Peterborough County—City Health Unit), 60 (Simcoe Muskoka District Health Unit), 27 (Brant County Health Unit), 34 (Haldimand-Norfolk Health Unit), 36 (Halton Regional Health Unit), 65 (Waterloo Health Unit), 66 (Wellington-Dufferin-Guelph Health Unit), 38 (Hastings and Prince Edward Counties Health Unit), 41 (Kingston, Frontenac and Lennox and Addington Health Unit), 43 (Leeds, Grenville and Lanark District Health Unit), 57 (Renfrew County and District Health Unit), 58 (The Eastern Ontario Health Unit), 26 (The District of Algoma Health Unit), 47 (North Bay Parry Sound District Health Unit), 49 (Northwestern Health Unit), 56 (Porcupine Health Unit), 61 (Sudbury and District Health Unit), 62 (Thunder Bay District Health Unit), 63 (Timiskaming Health Unit), 31 (Elgin-St. Thomas), 33 (Grey Bruce Health Unit), 39 (Huron County Health Unit), 40 (Chatham-Kent Health Unit), 42 (Lambton Health Unit), 54 (Perth District Health Unit), 75 (Southwestern Health Unit) |
| Syphilis testing | One or more syphilis serological screening tests positive in OLIS or LabWare categorized as yes/no or number of tests as 0, 1, 2, 3, and ≥4. Monthly rate of syphilis testing after dose 1 computed for each individual up until they either received their 2^nd^ dose, end of study period (October 31, 2023), or death. |
| Number of bacterial STIs | One or more bacterial STI (gonorrhea, chlamydia, or syphilis) diagnosis from iPHIS categorized as 0, 1, 2, 3, and ≥4. |
| HIV status | An ICES-specific HIV database was used to identify patients with HIV, based on 3 physician claims in 3 years with OHIP diagnostic codes: 042, 043, or 044.^1^ |
| HIV pre-exposure prophylaxis | HIV pre-exposure prophylaxis defined as a dispensation of tenofovir/emtricitabine recorded in the Ontario Drug Benefit (ODB) database, and excluding those prescribed other antiretrovirals within three months of the tenofovir/emtricitabine prescription.^4^ |
| History of receipt of non-MVA-BN vaccines in past one year | COVID-19 vaccines: The date of vaccine receipt from COVaXON.  Influenza vaccines:  Received in physician offices - OHIP feecodes: G590, G591, G592, Q130, Q590, Q690, Q691  Received in pharmacy - ODB billing with any of the following Drug Identification Numbers (DINs): 02420643, 02420783, 02432730, 02473283, 02445646, 02494248, 09857645, 09857646  Other vaccines received in physician offices:  OHIP feecodes: G840, G841, G842, G843, G844, G845, G846, G847, G848, G538, G539 |
| Neighborhood-level income quintiles | Household income quintile calculated at the disseminated area (DA) level was used for the neighborhood-level income.  A dissemination area (DA) is the smallest standard geographic area for which all census data are disseminated. A DA generally comprises approximately 400-700 people, but in densely populated cities may contain several thousand people. DAs cover all the territory of Canada.^3^ We assigned subjects to a DA using postal code, as recorded in the Registered Persons Database.  Calculated at the DA level using 2016 Census data by multiplying the median income (before-tax) by the number of households and dividing by the sum of single-person equivalent to obtain income per single person equivalent.^4^ For DAs where median income was unavailable, neighboring DAs were used to estimate income per single person equivalent. DA-based income quintiles were constructed separately for each census metropolitan area or census agglomeration (one or more adjacent municipalities integrated via commuting flows). DAs within each such area were ranked from the lowest average income per single-person equivalent to the highest, and DAs were assigned to five groups, such that each group contained approximately one-fifth the total in-scope population of each area. |
| Neighbourhood level visible minority quintiles | Those who self-identify as a visible minority. Calculated at the DA level using 2016 Census data. Ranked from lowest to highest and assigned to five groups, |
| Immigration status | Immigration application records for people who initially applied to land in Ontario. The data contains permanent residents' demographic information such as country of citizenship, level of education, mother tongue, and landing date. Categorized as: Refugees, immigration < 5 years, immigration 5-10 years, immigration >10 years, born in Canada or immigrated before 1985 (long-term residents). |
| Has a primary care physician | Whether patient is registered to a physician via the Client Agency Program Enrolment (CAPE) on index date. |
| Number of physician office visits | Any outpatient (office, home, or phone) OHIP visit/consult submitted by any physician via OHIP billing codes. |
| Moderately or severely immunocompromised | Includes Dialysis, Hematopoetic stem cell transplant recipient,  Solid organ transplant recipient, Receiving active treatment for solid tumour or hematologic malignancy, and/or Primary immunodeficiency. |

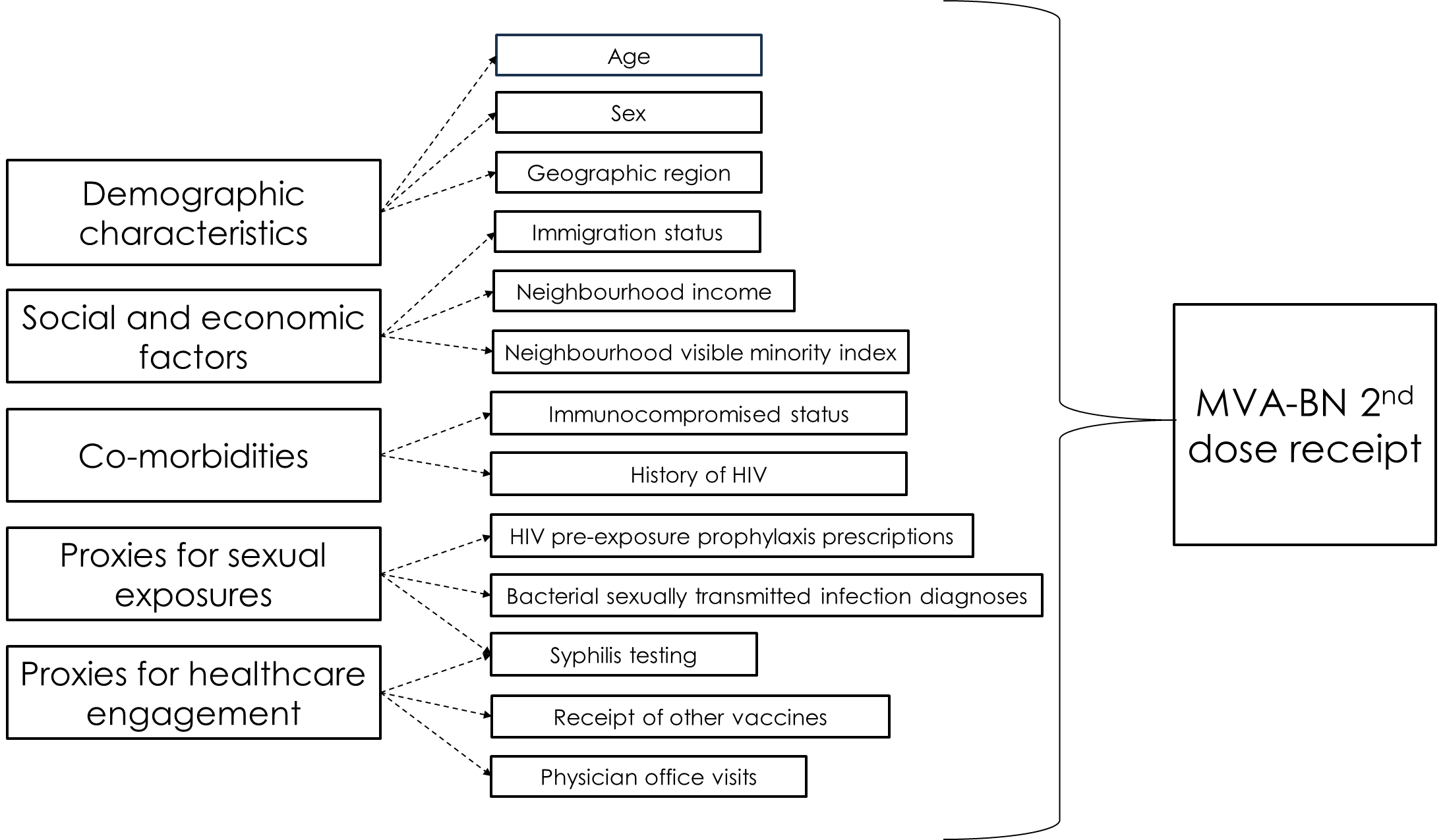

### **Figure S1.** Conceptual framework for associations explored between characteristics and MVA-BN dose 2 receipt.

Number of individauls

| Time between first and second dose (days) | Before September 30 |
| --- | --- |
| 1 | 1 |
| 2 | 1 |
| 6 | 1 |
| 13 | 1 |
| 19 | 1 |
| 22 | 2 |
| 23 | 1 |
| 24 | 1 |
| 25 | 5 |
| 26 | 7 |
| 27 | 2 |
| 28 | 31 |
| 29 | 21 |
| 30 | 16 |
| 31 | 7 |
| 32 | 16 |
| 33 | 19 |
| 34 | 10 |
| 35 | 24 |
| 36 | 12 |
| 37 | 9 |
| 38 | 17 |
| 39 | 10 |
| 40 | 11 |
| 41 | 11 |
| 42 | 25 |
| 43 | 10 |
| 44 | 5 |
| 45 | 14 |
| 46 | 11 |
| 47 | 9 |
| 48 | 20 |
| 49 | 21 |
| 50 | 21 |
| 51 | 13 |
| 52 | 7 |
| 53 | 14 |
| 54 | 7 |
| 55 | 18 |
| 56 | 39 |
| 57 | 14 |
| 58 | 14 |
| 59 | 27 |
| 60 | 11 |
| 61 | 21 |
| 62 | 27 |
| 63 | 50 |
| 64 | 21 |
| 65 | 21 |
| 66 | 19 |
| 67 | 10 |
| 68 | 19 |
| 69 | 24 |
| 70 | 54 |
| 71 | 37 |
| 72 | 27 |
| 73 | 17 |
| 74 | 26 |
| 75 | 41 |
| 76 | 41 |
| 77 | 57 |
| 78 | 35 |
| 79 | 31 |
| 80 | 30 |
| 81 | 44 |
| 82 | 48 |
| 83 | 44 |
| 84 | 71 |
| 85 | 47 |
| 86 | 25 |
| 87 | 42 |
| 88 | 38 |
| 89 | 25 |
| 90 | 47 |
| 91 | 81 |
| 92 | 41 |
| 93 | 42 |
| 94 | 28 |
| 95 | 34 |
| 96 | 39 |
| 97 | 55 |
| 98 | 65 |
| 99 | 60 |
| 100 | 43 |
| 101 | 48 |
| 102 | 56 |
| 103 | 82 |
| 104 | 109 |
| 105 | 171 |
| 106 | 116 |
| 107 | 115 |
| 108 | 74 |
| 109 | 86 |
| 110 | 104 |
| 111 | 140 |
| 112 | 210 |
| 113 | 144 |
| 114 | 139 |
| 115 | 99 |
| 116 | 104 |
| 117 | 174 |
| 118 | 136 |
| 119 | 186 |
| 120 | 130 |
| 121 | 133 |
| 122 | 98 |
| 123 | 80 |
| 124 | 83 |
| 125 | 82 |
| 126 | 93 |
| 127 | 69 |
| 128 | 71 |
| 129 | 76 |
| 130 | 44 |
| 131 | 51 |
| 132 | 64 |
| 133 | 74 |
| 134 | 37 |
| 135 | 41 |
| 136 | 36 |
| 137 | 28 |
| 138 | 45 |
| 139 | 41 |
| 140 | 56 |
| 141 | 48 |
| 142 | 26 |
| 143 | 30 |
| 144 | 25 |
| 145 | 21 |
| 146 | 40 |
| 147 | 58 |
| 148 | 26 |
| 149 | 27 |
| 150 | 27 |
| 151 | 23 |
| 152 | 24 |
| 153 | 30 |
| 154 | 34 |
| 155 | 19 |
| 156 | 26 |
| 157 | 19 |
| 158 | 20 |
| 159 | 22 |
| 160 | 28 |
| 161 | 46 |
| 162 | 20 |
| 163 | 17 |
| 164 | 20 |
| 165 | 19 |
| 166 | 16 |
| 167 | 23 |
| 168 | 18 |
| 169 | 19 |
| 170 | 25 |
| 171 | 14 |
| 172 | 13 |
| 173 | 20 |
| 174 | 16 |
| 175 | 36 |
| 176 | 28 |
| 177 | 31 |
| 178 | 15 |
| 179 | 25 |
| 180 | 17 |
| 181 | 26 |
| 182 | 32 |
| 183 | 32 |
| 184 | 19 |
| 185 | 18 |
| 186 | 15 |
| 187 | 22 |
| 188 | 20 |
| 189 | 24 |
| 190 | 10 |
| 191 | 18 |
| 192 | 11 |
| 193 | 20 |
| 194 | 16 |
| 195 | 23 |
| 196 | 28 |
| 197 | 30 |
| 198 | 23 |
| 199 | 16 |
| 200 | 14 |
| 201 | 13 |
| 202 | 15 |
| 203 | 25 |
| 204 | 22 |
| 205 | 21 |
| 206 | 23 |
| 207 | 11 |
| 208 | 16 |
| 209 | 11 |
| 210 | 32 |
| 211 | 22 |
| 212 | 26 |
| 213 | 21 |
| 214 | 23 |
| 215 | 19 |
| 216 | 27 |

| Time between first and second dose (days) | On/After September 30 |
| --- | --- |
| 14 | 1 |
| 20 | 1 |
| 21 | 1 |
| 23 | 2 |
| 25 | 1 |
| 27 | 10 |
| 28 | 191 |
| 29 | 60 |
| 30 | 53 |
| 31 | 46 |
| 32 | 39 |
| 33 | 54 |
| 34 | 30 |
| 35 | 128 |
| 36 | 30 |
| 37 | 29 |
| 38 | 18 |
| 39 | 21 |
| 40 | 14 |
| 41 | 20 |
| 42 | 76 |
| 43 | 14 |
| 44 | 16 |
| 45 | 9 |
| 46 | 6 |
| 47 | 7 |
| 48 | 15 |
| 49 | 32 |
| 50 | 24 |
| 51 | 14 |
| 52 | 7 |
| 53 | 10 |
| 54 | 12 |
| 55 | 19 |
| 56 | 27 |
| 57 | 9 |
| 58 | 4 |
| 59 | 10 |
| 60 | 6 |
| 61 | 7 |
| 62 | 10 |
| 63 | 26 |
| 64 | 7 |
| 65 | 6 |
| 66 | 4 |
| 67 | 15 |
| 68 | 4 |
| 69 | 6 |
| 70 | 25 |
| 72 | 5 |
| 73 | 2 |
| 74 | 1 |
| 75 | 2 |
| 76 | 6 |
| 77 | 13 |
| 78 | 2 |
| 79 | 4 |
| 80 | 1 |
| 81 | 1 |
| 82 | 4 |
| 83 | 5 |
| 84 | 16 |
| 85 | 7 |
| 86 | 3 |
| 87 | 3 |
| 88 | 1 |
| 89 | 1 |
| 90 | 3 |
| 91 | 11 |
| 92 | 6 |
| 93 | 5 |
| 94 | 2 |
| 95 | 1 |
| 96 | 4 |
| 97 | 2 |
| 98 | 8 |
| 99 | 6 |
| 100 | 4 |
| 101 | 1 |
| 102 | 4 |
| 103 | 1 |
| 104 | 4 |
| 105 | 6 |
| 106 | 1 |
| 107 | 2 |
| 108 | 2 |
| 109 | 1 |
| 110 | 4 |
| 111 | 2 |
| 112 | 5 |
| 113 | 6 |
| 115 | 1 |
| 116 | 1 |
| 117 | 2 |
| 118 | 3 |
| 119 | 3 |
| 120 | 2 |
| 121 | 1 |
| 122 | 1 |
| 123 | 2 |
| 124 | 2 |
| 125 | 2 |
| 126 | 5 |
| 128 | 4 |
| 129 | 1 |
| 130 | 1 |
| 131 | 3 |
| 132 | 2 |
| 133 | 3 |
| 136 | 1 |
| 137 | 1 |
| 138 | 3 |
| 139 | 1 |
| 140 | 5 |
| 141 | 2 |
| 143 | 1 |
| 144 | 3 |
| 145 | 1 |
| 146 | 3 |
| 147 | 2 |
| 148 | 1 |
| 149 | 1 |
| 152 | 4 |
| 153 | 2 |
| 154 | 5 |
| 155 | 1 |
| 156 | 2 |
| 158 | 2 |
| 159 | 1 |
| 160 | 2 |
| 161 | 2 |
| 162 | 5 |
| 163 | 1 |
| 164 | 1 |
| 165 | 1 |
| 166 | 1 |
| 167 | 1 |
| 168 | 1 |
| 169 | 2 |
| 170 | 2 |
| 171 | 4 |
| 174 | 1 |
| 175 | 3 |
| 176 | 4 |
| 179 | 2 |
| 180 | 1 |
| 182 | 4 |
| 183 | 5 |
| 185 | 1 |
| 189 | 2 |
| 194 | 1 |
| 196 | 1 |
| 197 | 2 |
| 200 | 1 |
| 202 | 1 |
| 203 | 2 |
| 205 | 2 |
| 207 | 1 |
| 210 | 1 |
| 211 | 1 |
| 212 | 3 |
| 214 | 1 |
| 215 | 3 |
| 217 | 1 |
| 218 | 1 |
| 219 | 2 |
| 220 | 1 |
| 222 | 1 |
| 223 | 1 |
| 229 | 1 |
| 231 | 1 |
| 232 | 1 |
| 234 | 2 |
| 235 | 2 |
| 236 | 1 |
| 238 | 1 |
| 240 | 1 |
| 241 | 1 |
| 244 | 1 |
| 245 | 1 |
| 252 | 1 |
| 255 | 1 |
| 260 | 2 |
| 263 | 1 |
| 264 | 1 |
| 265 | 1 |
| 287 | 1 |
| 290 | 1 |
| 294 | 2 |
| 301 | 1 |
| 303 | 1 |
| 309 | 1 |
| 315 | 1 |
| 320 | 2 |
| 323 | 1 |
| 328 | 1 |
| 331 | 1 |
| 332 | 1 |
| 338 | 1 |
| 341 | 1 |

### **Figure S2.** Time in days between first and second dose among individuals who had first dose before September 30, 2022 versus on/after September 30, 2022.
